# Auto-segmentation of thoraco-abdominal organs in pediatric dynamic MRI

**DOI:** 10.1101/2024.05.04.24306582

**Authors:** Yusuf Akhtar, Jayaram K. Udupa, Yubing Tong, Tiange Liu, Caiyun Wu, Rachel Kogan, Mostafa Al-noury, Mahdie Hosseini, Leihui Tong, Samarth Mannikeri, Dewey Odhner, Joseph M. Mcdonough, Carina Lott, Abigail Clark, Patrick J. Cahill, Jason B. Anari, Drew A. Torigian

## Abstract

**Purpose:** Analysis of the abnormal motion of thoraco-abdominal organs in respiratory disorders such as the Thoracic Insufficiency Syndrome (TIS) and scoliosis such as adolescent idiopathic scoliosis (AIS) or early onset scoliosis (EOS) can lead to better surgical plans. We can use healthy subjects to find out the normal architecture and motion of a rib cage and associated organs and attempt to modify the patient’s deformed anatomy to match to it. Dynamic magnetic resonance imaging (dMRI) is a practical and preferred imaging modality for capturing dynamic images of healthy pediatric subjects. In this paper, we propose an auto-segmentation set-up for the lungs, kidneys, liver, spleen, and thoraco-abdominal skin in these dMRI images which have their own challenges such as poor contrast, image non-standardness, and similarity in texture amongst gas, bone, and connective tissue at several inter-object interfaces.

**Methods:** The segmentation set-up has been implemented in two steps: recognition and delineation using two deep neural network (DL) architectures (say DL-R and DL-D) for the recognition step and delineation step, respectively. The encoder-decoder framework in DL-D utilizes features at four different resolution levels to counter the challenges involved in the segmentation. We have evaluated on dMRI sagittal acquisitions of 189 (near-)normal subjects. The spatial resolution in all dMRI acquisitions is 1.46 mm in a sagittal slice and 6.00 mm between sagittal slices. We utilized images of 89 (10) subjects at end inspiration for training (validation). For testing we experimented with three scenarios: utilizing (1) the images of 90 (=189-89-10) different (remaining) subjects at end inspiration for testing, (2) the images of the aforementioned 90 subjects at end expiration for testing, and (3) the images of the aforesaid 99 (=89+10) subjects but at end expiration for testing. In some situations, we can take advantage of already available ground truth (GT) of a subject at a particular respiratory phase to automatically segment the object in the image of the same subject at a different respiratory phase and then refining the segmentation to create the final GT. We anticipate that this process of creating GT would require minimal post hoc correction. In this spirit, we conducted separate experiments where we assume to have the ground truth of the test subjects at end expiration for scenario (1), end inspiration for (2), and end inspiration for (3).

**Results:** Amongst these three scenarios of testing, for the DL-R, we achieve a best average location error (LE) of about 1 voxel for the lungs, kidneys, and spleen and 1.5 voxels for the liver and the thoraco- abdominal skin. The standard deviation (SD) of LE is about 1 or 2 voxels. For the delineation approach, we achieve an average Dice coefficient (DC) of about 0.92 to 0.94 for the lungs, 0.82 for the kidneys, 0.90 for the liver, 0.81 for the spleen, and 0.93 for the thoraco-abdominal skin. The SD of DC is lower for the lungs, liver, and the thoraco-abdominal skin, and slightly higher for the spleen and kidneys.

**Conclusions:** Motivated by applications in surgical planning for disorders such as TIS, AIS, and EOS, we have shown an auto-segmentation system for thoraco-abdominal organs in dMRI acquisitions. This proposed setup copes with the challenges posed by low resolution, motion blur, inadequate contrast, and image intensity non-standardness quite well. We are in the process of testing its effectiveness on TIS patient dMRI data.

## 1. Introduction

The study of the motion of thoraco-abdominal organs can give better insights for treating patients with respiratory restrictive disorders such as Thoracic Insufficiency Syndrome (TIS) [1], adolescent idiopathic scoliosis (AIS) [2, 3], and early onset scoliosis (EOS) [4]. Consider for example, TIS, which is a pediatric disorder in which there is inability of the thorax to support normal respiration or lung growth, leading to complications. The constraints imposed by the abnormal osseous structures of TIS patients on the motion of other thoraco-abdominal organs such as the lungs, kidneys, liver, and spleen is currently not understood. Unfortunately, even in the case of normal subjects these motions are not well studied in pediatrics. As such, gathering a normative database consisting of dynamic images of healthy subjects during natural respiration and the analysis of the motion of these organs is essential to understand the deviation from normalcy of the architecture and motion of the organs in TIS patients. Such dynamic properties during respiration can be captured effectively via dynamic magnetic resonance imaging (dMRI) [1], which does not involve radiation exposure, does not require special patient maneuvers or breathing control, and can be implemented readily on MRI scanners available in the community. In this paper, we focus on the problem of auto-segmentation of the lungs, kidneys, liver, spleen, and thoraco- abdominal skin in dMRI sagittal acquisitions of healthy subjects which is an inevitable first step before carrying out motion analysis from dMRI.

We searched literature on dMRI images using the terms “dynamic MRI article”, “dynamic MRI segmentation article”, and “dynamic MRI thorax” in the Google search engine. The search with “dynamic MRI article” listed 2 articles, which presented cardiac motion [5] and musculoskeletal joint motion [6] from a clinical perspective only. The latter search “dynamic MRI segmentation article” listed 5 articles [7, 8, 9, 10, 11] which were related to segmentation of a single object of interest. For example, [9] dealt with segmentation of the blood vessels using a classical image processing approach. Another article [11] presented a method to segment the skin from axial slices of the breast. Three articles [12, 13, 14], which were listed with “dynamic MRI thorax”, utilized only manual segmentations of diaphragm or chest wall excursions for measuring relevant physiological parameters. There exist two works [15, 16] which deal with the segmentation of the lungs in dMRI images. To the best of our knowledge, methods dealing with multi-organ (>2: lungs, liver, spleen, kidneys, and thoraco-abdominal skin) automatic segmentation in dMRI acquisitions, especially of the thorax/abdomen, do not exist. We will discuss the related works [15, 16] and other articles referenced in this paragraph in further detail in the next section.

Static MRI images have lower spatial resolution compared to computed tomography (CT) images, and are prone to various challenges. These problems also occur in dMRI acquisitions even more severely: (1) different meaning of gray-level intensities for the same object for the same subject across different acquisitions, and different meaning of gray-level intensities for the same object across different subjects, (1) (2) poor contrast amongst objects, (3) low signal-to-noise ratio, (4) motion blur, (5) low spatial resolution, and (6) similarity in intensity and texture amongst gas, bone, and connective tissues at several inter-object interfaces. These issues make multi-organ segmentation in dMRI images very challenging (Figure 1). To further elaborate (6) above, the peripheral region of the lungs can be confused with the surrounding connective tissue. The posterior portion of the liver can be confused with the stomach and the spleen while the inferior portion of the liver can be confused with the gastro-intestinal tract. The posterior portion of the kidneys can be confused with the muscles connected to the spine while the anterior portion and the inferior portion of the spleen could be confused with the stomach and the left kidney, respectively.

**Figure 1:**
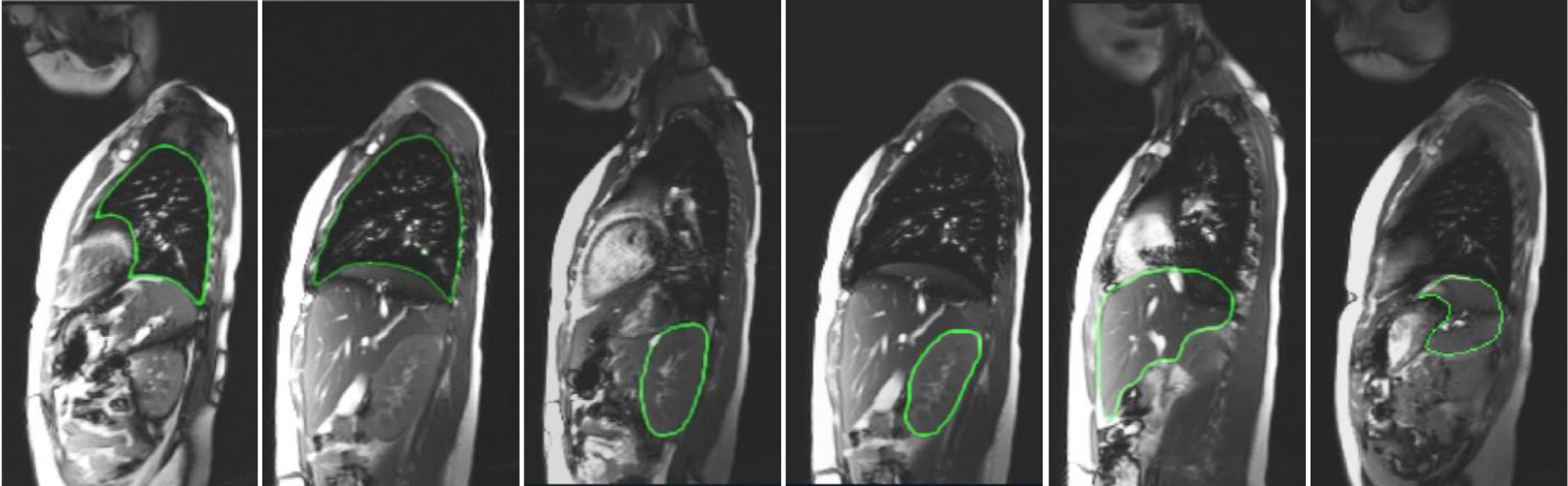
Representative sagittal bright-blood dMRI slices at end-expiratory phase (obtained with 4D construction [17] from a dMRI acquisition) through thorax and abdomen of a normal subject with true boundary delineations for (left to right) left lung, right lung, left kidney, right kidney, liver, and spleen.

Dynamic MRI acquisitions are inherently four dimensional with the dimensions being space (in three dimensions) and time. In our dMRI acquisition, a sagittal slice MR image at a fixed location is first acquired continuously for a specified duration (typically over 10 respiratory cycles) while the subject is breathing naturally, and then the next sagittal slice is captured for the next specified duration, and so on until the right to left width of the entire thoraco-abdominal region is fully covered. To segment the thoraco-abdominal organs, we first perform a 4D construction of the body region image representing the dynamic body region over one respiratory cycle via an optical flux strategy [17], and then segment the 3D organs (see Figure 2) in the 3D images corresponding to specified respiratory phases such as the end-inspiration (EI) and end-expiration (EE) time point phases. Our contribution in this paper is that we present a novel and unique system to address the problem of multi-organ segmentation from dMRI acquisitions of the thoraco-abdominal region.

**Figure 2:**
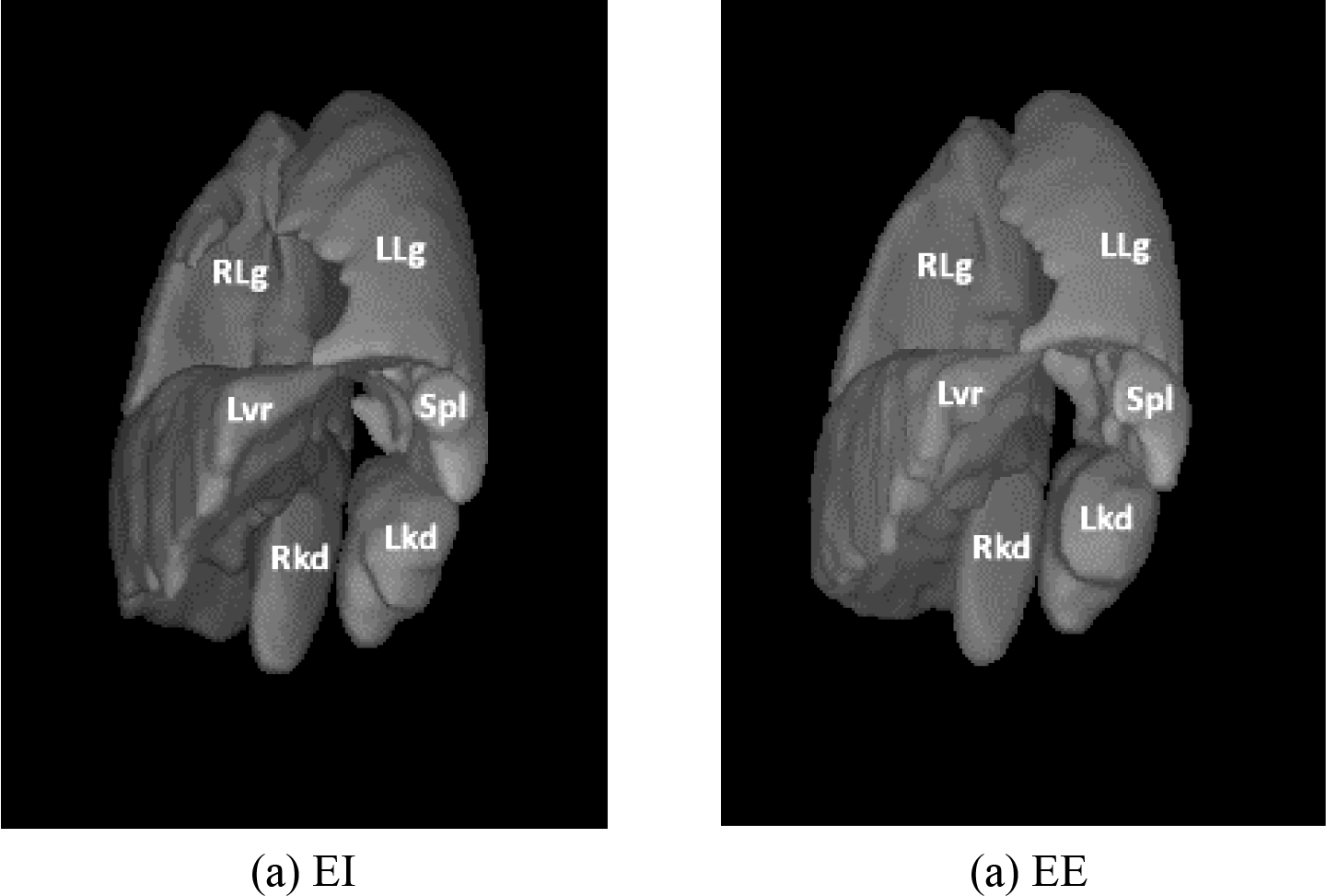
Three-dimensional rendering of the ground truth segmentation of the left lung (LLg), right lung (RLg), left kidney (Lkd), right kidney (Rkd), liver (Lvr) and spleen (Spl) for a normal subject at (a) end inspiration (EI) and (b) end expiration (EE).

A preliminary version of this work was presented at the SPIE 2023 Medical Imaging Conference whose proceedings contained a very abbreviated version of this work. The conference paper [18] differs from the current paper in the following manner.

1. The 3D images at EI only were utilized in [18]. The current paper utilizes images at multiple (two (EI and EE) and greater than two) respiratory phases.
2. The conference paper focused on the large organs – left lung and right lung only. The current paper includes the more challenging left kidney, right kidney, liver, spleen, and thoraco- abdominal skin as well.
3. This paper includes further expansions. For example, we show how an additional information from one respiratory phase can be considered for the delineation of the object in the image of the same test subject in a different respiratory phase.
4. Our experimental evaluation involves significant expansion over the conference paper, with a much larger data set (189 vs. 95 in the conference paper) and additional experiments involving repeated scans to show the consistency of performance of the proposed method.

## 2. Related Work

The articles [5, 6, 7, 8, 9, 10, 11, 12, 13, 14, 19] were found to be relevant to dMRI acquisitions for medical imaging applications. These can be grouped into three categories. A: articles which review the viability of dMRI for specific clinical applications. B: articles which discuss dMRI for measurement of physiological parameters with the help of segmentation algorithms. C: articles which discuss dMRI in the context of clinical applications but with manual analysis in these dMRI images. D: other articles such as those discussing image reconstruction in dMRI. We give a brief overview of the articles in these categories.

The articles [5, 6] in Category A are not related to segmentation of objects in dMRI images. Specifically, the authors in [5] tried to show how dynamic contrast-enhanced MRI (DCE-MRI) is an attractive imaging modality for measuring peripheral perfusion, other diverse microvascular parameters such as vessel permeability and fluid volume fractions, and the actual tissue perfusion. The authors in [6] discussed techniques of using dMRI for evaluating joint motion.

The articles [7, 8, 9, 10, 11] in category B, deal with segmentation of a single object of interest in dMRI images. The articles [8, 9, 10, 11] did not elucidate their segmentation methods. The authors in [7] used dMRI to assess pelvic organ prolapse with segmentation of the vertebral shape. Specifically, they segmented the sacral curve using a curve fitting procedure, and utilized an adaptive shortest path algorithm that enhances edge detection and linking. They report discerning the sacral curve from dMRI in 91% of the images. Since we are dealing with volumetric objects compared to linear objects, we cannot directly apply their method to segment the lungs, kidneys, liver, spleen, or thoraco-abdominal skin. The authors in [8] demonstrated the feasibility of quantitative cerebral blood flow (CBF) measurements during supine bicycling exercise with pseudo-continuous arterial spin labeling MRI at 3T. The authors in [9] utilized classical segmentation algorithms to segment the foci which represent tumor in the prostate gland in the dMRI image. The authors in [10] tried to indirectly measure local changes in CBF, blood volume, and blood oxygenation from neuronal activity via segmentation in the dMRI image. Lastly, the authors in [11] proposed a method to segment and remove the skin from the dMRI image of the breast to improve the clarity of the breast tissue in the dMRI image for further diagnosis. We reiterate that the articles cited in this paragraph deal with the segmentation of a single object of interest in dMRI images, they do not deal with dynamic or moving thoraco-abdominal organs, and some of the dMRI images pertain to studying the kinetics of the contrast agent and not the dynamic motion of organs.

The article [12] uses dMRI images to measure the chest wall and diaphragm excursions in normal subjects. However, they utilize the manual segmentations of the lungs and the chest wall for evaluating the relevant excursion parameters. In [13] diaphragm function is analyzed from their manual segmentations in dMRI images. In [14] the benign and malignant tumor lesions in lungs are manually segmented. These manual segmentations are utilized directly to establish their characteristics in dMRI acquisitions.

We found one article [19] that belongs to category D. The authors in [19] discuss a novel approach for reconstructing the dMRI image quickly from k-space and spatial prior knowledge via a multi-supervised network which they call “DIMENSION”. However, the article [19] does not deal with segmentation.

The authors of [15] had proposed a deep neural network for segmentation of the lungs from dMRI sagittal acquisitions of (near-)normal pediatric subjects using a U-Net architecture. The article [16] uses atlas based segmentation approaches for the lungs in dMRI images. In the current paper, our segmentation approach uses the neural networks in [20, 21] where the encoder-decoder architecture is enhanced with different modules such as the Path Aggregation Network (PAN) [22] and Dual Attention Network (DAN) [23]. We adopted this enhanced architecture as we are also dealing with segmentation of the left kidney, right kidney, liver, and spleen, which are more challenging (given poor contrast and inconsistent intensity meanings) to handle than the segmentation of the lungs. From the above discussion, we conclude that except for [15, 16], the problem of multi-organ (say greater than 2 organs) segmentation in the thoraco- abdominal region of dMRI acquisitions has not been addressed before.

## 3. Methods

### 3.0 Data acquisition and pre-processing

*dMRI scans*: The dMRI scan data were acquired from 189, 6-20 year-old, healthy children under an ongoing prospective research study protocol approved by the Institutional Review Board at the Children’s Hospital of Philadelphia (CHOP) and University of Pennsylvania, along with Health Insurance Portability and Accountability Act waiver. We excluded scans with significant body movement during scanning or with obvious image artifacts. The thoracic dMRI protocol includes a 3T MRI scanner (Verio, Siemens, Erlangen, Germany) using a True-FISP sequence with acquisition and reconstruction parameters of TR=3.82 ms, TE=1.91 ms, flip angle 76 degrees, bandwidth 258 Hz, 320×320 matrix, and voxel size ∼1×1×6 mm^3^. For each sagittal location across the thorax, 80 image slices were obtained over several tidal breathing cycles at ∼480 ms/slice. On average, 35 sagittal locations across the chest were imaged. Therefore, a total of 2800 (35 × 80) 2D MRI slices were acquired per subject.

*4D construction*: Given the dMRI scan for each subject, a small set of 175-320 slices representing one 4D volume over one respiratory cycle is selected from the 2800 2D free-breathing dMRI image slices using an optical flux-based optimization method to represent the dynamic thorax of the subject [17].

*Image intensity standardization*: MRI signal intensities in the 4D constructed image are standardized [24] to a standard intensity scale to facilitate MRI segmentation and analysis. Intensity standardization enables voxel intensity values to have similar numeric meaning for each type of tissue within the same subject, across subjects, in repeat scans on the same scanner, and across different scanners [24, 25].

*Creating ground truth segmentations*: Following the principles outlined in [26], we created clinically meaningful and computationally feasible definitions of the thoraco-abdominal body region and objects considered in this application to make the models anatomically specific and minimize inter-tracer variability while creating the ground truth of the objects. We define the thoraco-abdominal body region considered in this application as extending from 15 mm superior to the lung apices to the inferior aspect of the kidneys. Similarly, each object was defined in terms of which substructures are to be included/excluded. A board-certified radiologist with more than twenty-five years of experience (Torigian) trained students, post-doctoral fellows, engineers, and medical interns (Kogan, Tong L, Mannikeri, Akhtar, Wu, Al-noury) for anatomic and dMRI radiological appearance of the relevant structures. Following training, the 7 organs of our focus in the 189 dMRI acquisitions were all segmented manually by the above individuals through use of the open-source software CAVASS [24] in the EE and EI time points of the respiratory cycle. This yielded a total of 2,646 (=189x7x2) 3D object samples for our cohort.

### 3.1 Segmentation: DL-R [20]

We perform segmentation in two steps: a recognition step and a delineation step (Figure 3). In recognition, we try to obtain a rough idea of the location of the object of interest in the unseen image with the help of bounding boxes. In delineation, the approach marks the outline of the object of interest within the bounding box. We have utilized deep learning recognition (DL-R) and deep learning delineation (DL-D) networks for recognition and delineation. We now give a description of DL-R in the rest of this section and a description of DL-D in section 3.2.

**Figure 3:**
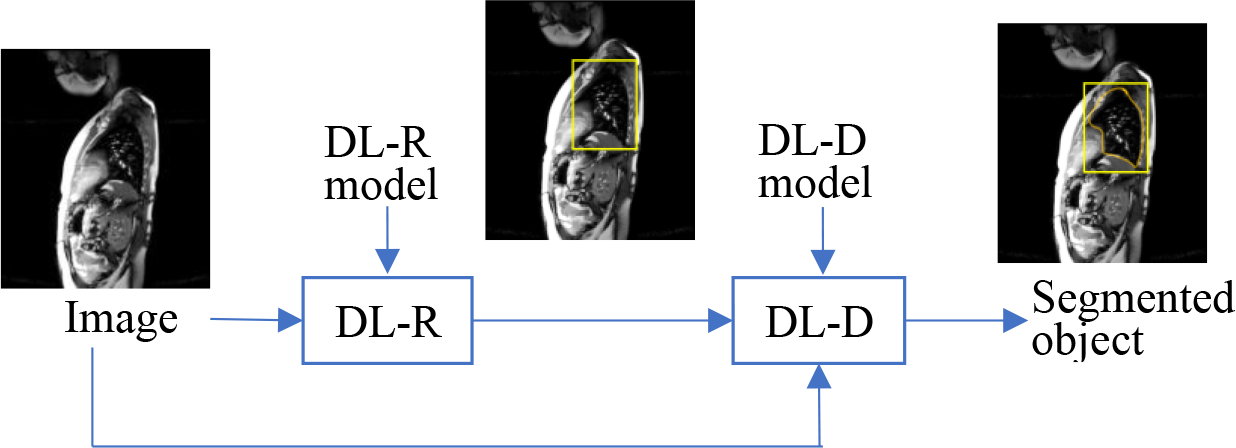
Illustration of our segmentation pipeline. Deep learning recognition (DL-R) module used for the recognition step. Deep learning delineation (DL-D) module utilized for the delineation step .

The DL-R module consists of three types of networks: backbone network, neck network, and head network. The backbone network is based on ResNet [27] and DenseNet [28]. The input to the backbone network is a 3-channel image which is obtained by mapping each pixel intensity value in a sagittal slice to three intensity values using three pre-defined intensity intervals. The backbone network uses pre-trained model weights of ResNet and DenseNet. From the last four convolutional layers of the backbone network, four feature maps (C2, C3, C4, and C5) are taken using strides of 4, 8, 16, and 32 pixels, respectively. The map C2 captures lower-level textural information compared to C3, C4, and C5. The map C5 captures high level contextual information from the 3-channel input image. These feature maps (C2, C3, C4, and C5) are taken as 4 separate inputs (channels) to the neck network.

The neck network is based on PAN [22] and DAN [23] architectures. The PAN architecture creates maps referred to by Q4, Q5, and Q6 by merging feature maps C2, C3, C4, and C5 using bottom-up connections, top-down connections, and lateral connections. The DAN architecture is used to create prediction maps which contain the information dependency across the spatial dimensions and the channel dimensions of the maps Q4, Q5, and Q6. The maps Q4, Q5, Q6 and the prediction maps are taken as input to the head network.

The head network recognizes the non-sparse organs with the maps Q4, Q5, and Q6 by associating them with anchor sizes 32 x 32, 64 x 64, and 128 x 128, respectively. This recognition is further refined by utilizing the prediction maps and the anchors with the help of convolutional layers. The output of DL-R is a bounding box in those sagittal slices which are identified to contain the objects of interest. These bounding boxes come from the head network.

The presence of bifurcations of blood vessels in the liver or the appearance of chambers of the heart in the dMRI image can help in locating our organs of interest. Such signatures could also be inconspicuous to the naked eye if they are present at a local scale. The feature maps such as C2 that extracts low-level textural information is integrated with feature maps dealing with high-level contextual information (say from C5) in the neck network. This makes the usage of the DL-R approach appealing for our multi-organ segmentation problem, as it integrates different types of information at varying scales in its design.

For training and testing the DL-R model, the images are intensity standardized [25]. This step of intensity standardization is necessary because intensity values of an object of interest have different meanings in different dMRI acquisitions. Intensity standardization yields a tissue-specific numeric meaning for an object of interest in the transformed image, which helps in utilizing recognition algorithms effectively.

The DL-R model takes three thresholds as input for transforming a sagittal slice to the 3 (color)-channel (2D) image (refer the second paragraph of this subsection). The motivation behind creating this 3-channel image is to roughly visually depict different compositions of the body region based on intensity values alone. We observe that gas and cortical bone are very dark in appearance on bright-blood MRI images, as compared to soft tissues of the skeletal muscles and visceral organs which are somewhat brighter in appearance, and the fat, cardiac chambers, and blood vessels which appear very bright. We have chosen the first color channel to represent low intensity objects, the second color channel to represent medium intensity objects, and the third color channel to represent high intensity objects.

For each color channel, two thresholds (an upper threshold and a lower threshold) are chosen to roughly contain the intensities of interest. Based on our visual inspection of the histograms of the pixel intensity values of the objects of interest (left lung, right lung, left kidney, right kidney, liver, and spleen) in the intensity standardized images, we have chosen the three thresholds as 150 units, 750 units, and 1500 units. For example, consider a channel which uses a lower threshold (L) and an upper threshold (U). If the pixel value in the intensity standardized image is *y*, we transform *y* to 0 if it is less than L. If *y* lies between L and U, it is transformed to 255*(*y*-L)/(U-L). If *y* is greater than U, it is transformed to 255. For the first channel, L is 0 unit and U is 150 units. For the second channel, L is 150 units and U is 750 units. For the third channel, L is 750 units and U is 1500 units.

The DL-R module is optimized using an Adam optimizer with a learning rate of 0.00001. The Focal Loss [29] function is utilized for optimization of the DL-R module.

### 3.2 DL-D [21]

This module utilizes a network called ABCNet [21], which was originally designed to delineate the different types of body tissues: subcutaneous adipose tissue, visceral adipose tissue, skeletal muscle tissue, and skeletal tissue from low dose axial CT images of the body torso. The design of ABCNet is similar to an encoder-decoder architecture.

The fundamental unit of ABCNet is referred to by BasicConv which is comprised of four modules in succession: concatenation, batch normalization, activation, and convolution. Bottleneck is a special case of BasicConv with a convolutional kernel of 1x1x1. There are four DenseBlocks [28] used in the encoder-decoder architecture of ABCNet. The deeper the DenseBlock, the more high-level information it extracts from the input image. Each DenseBlock of ABCNet is composed of Dense Layers, which are themselves composed of Bottleneck and a BasicConv with a kernel size of 3x3x3 in succession. The bottleneck, because of its lower convolutional kernel size, keeps the number of parameters less and simultaneously acts as a feature extractor through the normalization and activation functions of its BasicConv architecture.

The ABCNet model uses a Dice coefficient-based loss function for training its model and selects patches randomly from within and slightly around the ground truth in the images of the seen dataset during training. During testing, the patches are selected from within and slightly around the bounding box (from the recognition step) in the images of the unseen dataset. The output of ABCNet is the prediction map of the object from the decoder. This prediction map is binarized using a threshold to yield the final segmentation of the object. Unlike existing encoder-decoder architectures (DeepMedic [30], Dense V-Net [31], V-Net [32], and 3D U-Net [33]), which have typically 12 or 31 layers and 1 million or 80 million parameters, ABCNet has 118 layers with only 1.4 million parameters. The usage of ABCNet is thus attractive because of its deeper architecture with a lower number of parameters.

We have used intensity normalization on the intensity standardized images before using them for training and testing the DL-D module. For intensity normalization, the Z-score method has been utilized. The Z-score utilizes the mean (and standard deviation) of the standardized pixel values belonging to the object over all images in the training set. Let this mean (standard deviation) be denoted by μ (σ). A pixel value x in an image of the training set or in the test set is transformed to a new value y by the relation y=(x-μ)/σ.

The patch size which is an input to the DL-D module is chosen as 72 x 72 x 24 voxels for large organs such as the left lung, right lung, liver, and thoraco-abdominal skin, and as 72 x 72 x 16 voxels for smaller organs such as the spleen, left kidney, and right kidney. Smaller patches lead to a reduction in the delineation accuracy of DL-D. Larger patches require a large amount of memory, sometimes exceeding the workstation’s memory capacity and exponentially increasing the time required for training the DL-R and DL-D models.

The DL-D module is trained for 50 epochs with 200 steps per epoch. A batch size of 4 is utilized for a mini-batch gradient descent for optimization. The initial learning rate is set to 0.01, which is reduced further to 0.00001 by the cosine annealing strategy.

## 4. Experiments and Results

As summarized in Table 1, we have conducted 9 experiments (Exp. 0–Exp. 8) to analyze the behavior of the whole pipeline. We particularly focus on the 3D images corresponding to EE and EI since they are critical for analyzing lung tidal volumes.

**Table 1:**
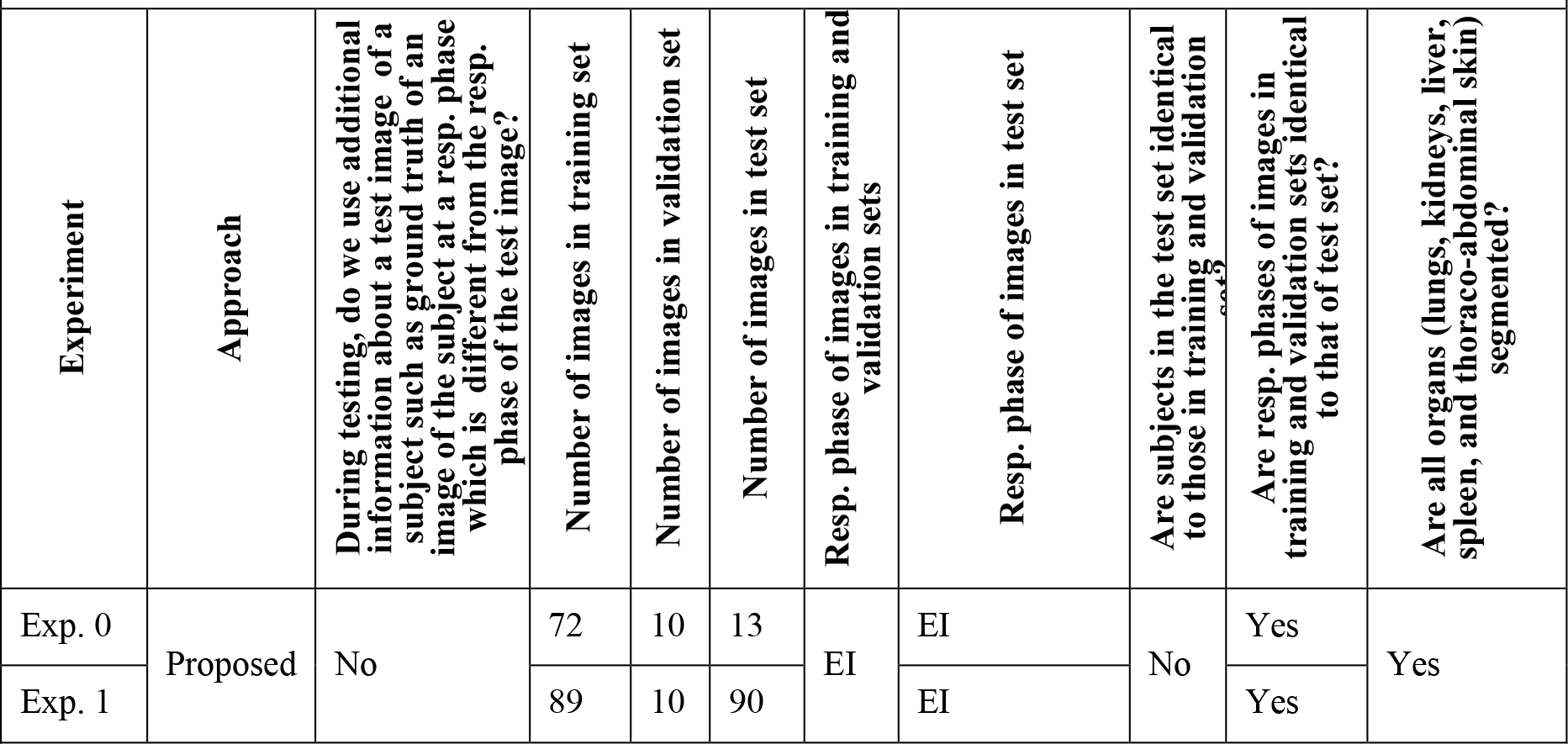
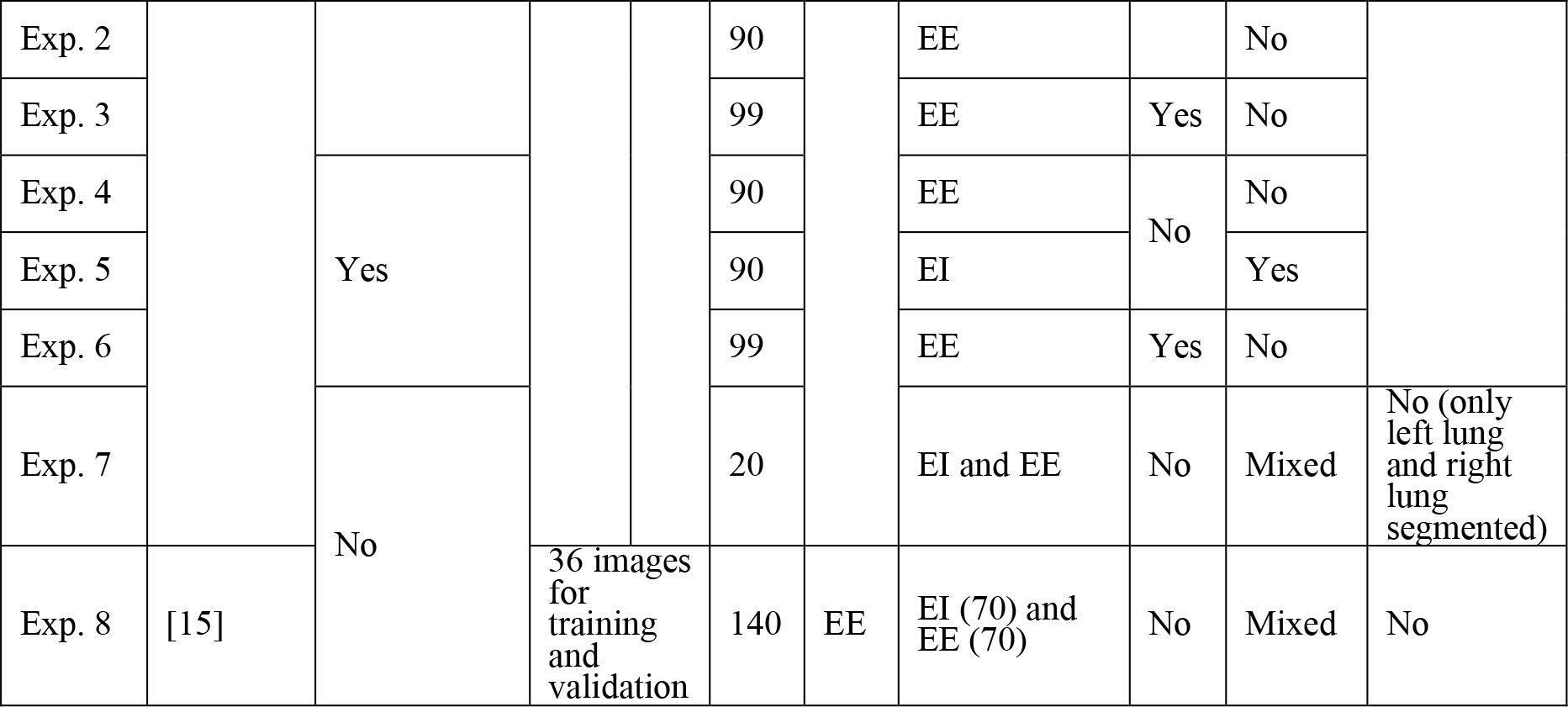
Summary of related information about the 9 experiments presented in this paper.

### 4.1. Exp. 0: Experiment which utilizes (3D) images at a single respiratory phase (EI) with data augmentation

In this experiment, we explored different data augmentation techniques using reflection of the dMRI images across a 2D plane (sagittal, transverse, or coronal) or reflection across a particular combination of the 2D planes. We examine each object and see what reflection mode makes anatomic sense for that object and examine the delineation output by DL-D for all methods of data augmentation. In Table 2, “0”, “1”, and “2” mean reflection is made across the transverse, coronal, or sagittal plane, respectively, and a serial listing of these numbers indicates a series of reflections (e.g., “01” means that reflection is first made across the transverse plane then across the coronal plane). The last column indicates no reflection.

**Table 2:** Mean (1^st^ value) and standard deviation (SD, 2^nd^ value) of DC over the tested data sets are listed for the 7 organs for the different data augmentation strategies. “0”, “1”, “2” mean reflection about the transverse, coronal, and sagittal planes, respectively. Multiple digits indicate reflection involving multiple planes. For example, “01” means reflection is first made across the transverse plane then across the coronal plane.

| Organ | "0" | "1" | "2" | "01" | "12" | "02" | "012" | None |
| --- | --- | --- | --- | --- | --- | --- | --- | --- |
| Left lung | 0.947<br>$\pm 0.021$ | 0.944<br>$\pm 0.013$ | 0.911<br>$\pm 0.085$ | 0.947<br>$\pm 0.024$ | 0.938<br>$\pm 0.033$ | 0.942<br>$\pm 0.028$ | 0.905<br>$\pm 0.052$ | **0.948**<br>**$\pm 0.021$** |
| Right lung | 0.945<br>$\pm 0.019$ | 0.956<br>$\pm 0.013$ | 0.954<br>$\pm 0.024$ | 0.907<br>$\pm 0.047$ | 0.933<br>$\pm 0.032$ | 0.946<br>$\pm 0.021$ | 0.942<br>$\pm 0.032$ | **0.958**<br>**$\pm 0.008$** |
| Left kidney | 0.805<br>$\pm 0.092$ | 0.772<br>$\pm 0.142$ | 0.799<br>$\pm 0.095$ | 0.785<br>$\pm 0.111$ | 0.840<br>$\pm 0.029$ | **0.855**<br>**$\pm 0.029$** | 0.774<br>$\pm 0.210$ | 0.820<br>$\pm 0.071$ |
| Right kidney | **0.871**<br>**$\pm 0.048$** | 0.806<br>$\pm 0.126$ | 0.793<br>$\pm 0.238$ | 0.685<br>$\pm 0.314$ | 0.780<br>$\pm 0.192$ | 0.781<br>$\pm 0.239$ | 0.839<br>$\pm 0.061$ | 0.855<br>$\pm 0.089$ |
| Liver | 0.865<br>$\pm 0.052$ | 0.856<br>$\pm 0.044$ | 0.823<br>$\pm 0.056$ | 0.866<br>$\pm 0.070$ | 0.895<br>$\pm 0.023$ | **0.911**<br>**$\pm 0.019$** | 0.893<br>$\pm 0.029$ | 0.891<br>$\pm 0.026$ |
| Spleen | 0.792<br>$\pm 0.109$ | **0.824**<br>**$\pm 0.053$** | 0.818<br>$\pm 0.069$ | 0.802<br>$\pm 0.077$ | 0.783<br>$\pm 0.102$ | 0.811<br>$\pm 0.066$ | 0.803<br>$\pm 0.067$ | 0.791<br>$\pm 0.089$ |
| Thoraco-abdominal skin | **0.911**<br>**$\pm 0.025$** | 0.896<br>$\pm 0.032$ | 0.906<br>$\pm 0.031$ | 0.893<br>$\pm 0.020$ | 0.888<br>$\pm 0.032$ | 0.902<br>$\pm 0.032$ | 0.887<br>$\pm 0.033$ | 0.908<br>$\pm 0.027$ |

dMRI images of 72, 10, and 13 subjects at EI were used for training, validation, and testing the DL-D module. In Table 2, the average Dice coefficient (*DC*) indicates that for each of the 7 organs, a particular method of reflection is optimal. However, such data augmentation techniques are not meaningful in the context of our thoraco-abdominal organ segmentation problem as the meaning behind the appearance of the organs in the reflected image changes with respect to the meaning of the appearance of the organs in the original image. The results of the last column of Table 2 with no reflection show excellent *DC* for most of the organs suggesting that this augmentation method is not useful.

### 4.2 Exp. 1-3: Experiments which utilize (3D) images at two respiratory phases (EI and EE)

The creation of ground truth (GT) in dMRI images is labor intensive and is impractical for all respiratory phases. Thus, if we can create GT or close to GT (which requires minimal post hoc correction rather than tracing from scratch) through our pipeline, it will be useful. We evidently need to train our networks for obtaining the DL-R and DL-D models which would be utilized for creating the GT. Certain scenarios during these training and testing processes would arise as discussed below.

We can use two parameters to describe the images in the test set or training set. The first parameter is the subject (PSub) to which an image belongs. The second parameter (PResp) is the respiratory phase to which the image belongs. Based on which of PSub and PResp are identical or different between the training and test set, there can be four combinations of training and testing. We exclude that combination where both PSub and PResp are identical in the training and test sets. The other three combinations of experimentations constitute Exp. 1, 2, and 3 in Table 1.

We explore three scenarios F, G and H (see Table 3). In scenario F, PResp is identical between training and test sets but PSub is different. In G, Presp and PSub are both different between training and test sets. In H, PResp is different but PSub is identical between training and test sets. We utilize 89 images at EI for training and 10 images at EI for validation in F, G, and H. For testing, we utilize in F: the images of 90 (=189-89-10) different (remaining) subjects at EI, G: the images of the aforementioned 90 subjects at EE, and H: the images of the aforesaid 99 (=89+10) subjects but at EE.

**Table 3:** Partitioning of 189 subjects for the evaluation of DL-R and DL-D modules for Exp. 1-3.

| Number of subjects | Respiratory phase of image | Purpose | Scenario | PResp | PSub |
| --- | --- | --- | --- | --- | --- |
| 99 | EI | Training<br>(89) and<br>Validation<br>(10) | F, G, and<br>H | --- | --- |
|  | EE |  | H | Different | Same |
| 90 | EI | Testing | F | Same | Different |
|  | EE |  | G | Different | Different |

In Exp. 1-3, we check scenarios F-H, respectively, and the results are summarized in Table 4 for DL- R and Tables 5-6 for DL-D including Dice coefficient (*DC)* and mean-Hausdorff distance (mean-*HD*). The recognition error for DL-R is expressed in terms of location error *LE* which is defined as the distance between the centroids of the bounding box from the recognition step and the tight-fitting bounding box around the true object (ground truth). Note that in Experiment 3, an image in the training set and an image in the test set can belong to the same subject dMRI acquisition. However, these two images (in the preceding statement) belong to different respiratory phases (as EE for the test set and as EI for the training set).

**Table 4:** Recognition location error (mean±SD: LE) in mm of DL-R on images at end inspiration (EI) and end expiration (EE) for experiments Exp. 1-3. The least LE under each column is highlighted.

| Test | Left lung | Right lung | Left kidney | Right kidney | Liver | Spleen | Thoraco-abdominal skin |
| --- | --- | --- | --- | --- | --- | --- | --- |
| Exp. 1<br>(F: 90<br>EI) | 5.69<br>$\pm 3.23$ | 6.31<br>$\pm 5.81$ | 4.66<br>$\pm 2.50$ | 5.16<br>$\pm 8.12$ | 7.69<br>$\pm 6.09$ | 5.50<br>$\pm 3.51$ | 7.18<br>$\pm 4.22$ |
| Exp. 2<br>(G: 90<br>EE) | 5.35<br>$\pm 3.38$ | 5.20<br>$\pm 2.95$ | 4.15<br>$\pm 2.45$ | 4.41<br>$\pm 2.80$ | 10.21<br>$\pm 8.27$ | 5.83<br>$\pm 3.52$ | 7.11<br>$\pm 4.25$ |
| Exp. 3<br>(H: 99<br>EE) | **4.73**<br>**$\pm 3.44$** | **4.90**<br>**$\pm 2.63$** | **3.79**<br>**$\pm 2.16$** | **3.70**<br>**$\pm 1.93$** | **6.94**<br>**$\pm 5.76$** | **4.31**<br>**$\pm 2.17$** | **4.09**<br>**$\pm 2.85$** |

**Table 5:** Delineation results (mean±SD: DC) of DL-D on images at end inspiration (EI) and end expiration (EE) for three experiments. The highest average Dice coefficient under a column has been highlighted. For further details, please refer subsection 4.2.

| Test | Left lung | Right lung | Left kidney | Right kidney | Liver | Spleen | Thoraco-abdominal skin |
| --- | --- | --- | --- | --- | --- | --- | --- |
| Exp. 1 (F: 90 EI) | 0.936<br>$\pm 0.018$ | 0.930<br>$\pm 0.039$ | 0.808<br>$\pm 0.094$ | 0.828<br>$\pm 0.072$ | 0.896<br>$\pm 0.053$ | 0.807<br>$\pm 0.075$ | 0.929<br>$\pm 0.029$ |
| Exp. 2 (G: 90 EE) | 0.924<br>$\pm 0.033$ | 0.923<br>$\pm 0.041$ | 0.811<br>$\pm 0.069$ | 0.833<br>$\pm 0.082$ | 0.899<br>$\pm 0.053$ | 0.826<br>$\pm 0.050$ | 0.925<br>$\pm 0.028$ |
| Exp. 3 (H: 99 EE) | **0.943**<br>**$\pm 0.026$** | **0.939**<br>**$\pm 0.028$** | **0.833**<br>**$\pm 0.056$** | **0.843**<br>**$\pm 0.077$** | **0.909**<br>**$\pm 0.023$** | **0.830**<br>**$\pm 0.057$** | **0.937**<br>**$\pm 0.024$** |

**Table 6:**
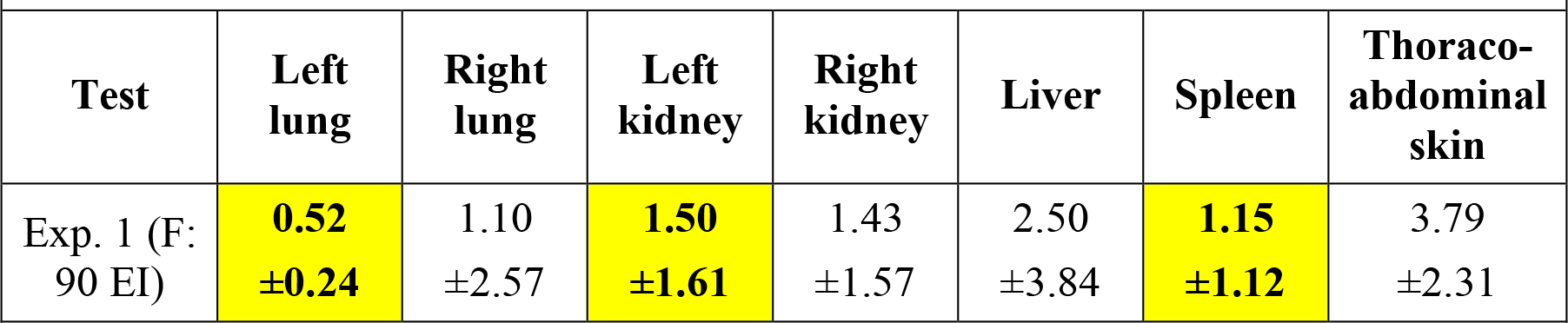
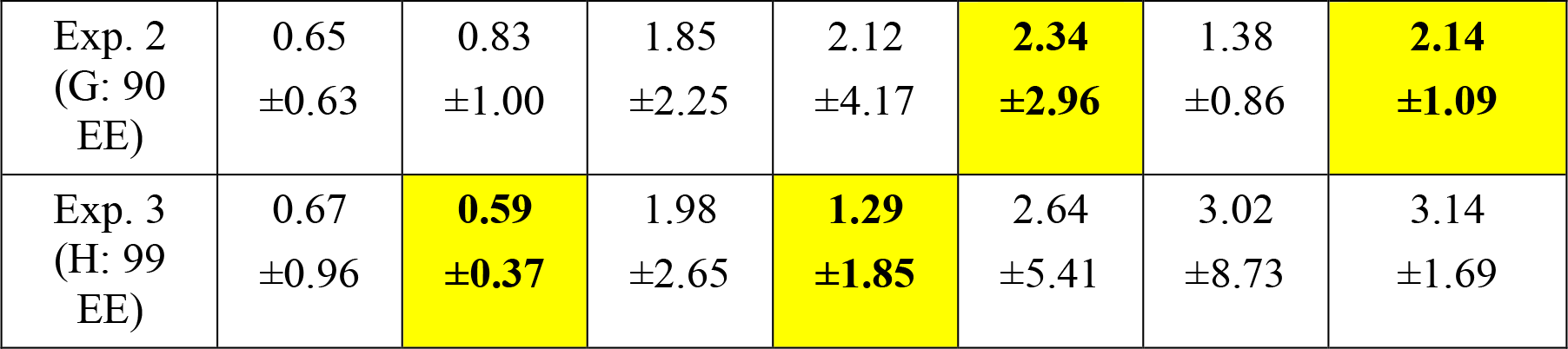
Performance of DL-D in terms of (mean±SD) mean-HD in units of mm for three experiments. The least value under a column has been highlighted. For further details, please refer subsection 4.2.

We notice from the results in Table 4 that out of the three experiments, Experiment 3 has the least average location error for all seven organs. This observation aligns with our intuition that if we have additional information about the test image (such as the ground truth in the image of a test subject at a particular respiratory phase), and use it in training DL-R, we will obtain better recognition results in the image of the same subject at a different respiratory phase.

Amongst Experiment 1, Experiment 2, and Experiment 3, we notice from the results in Table 5 and Table 6 that Experiment 3 fares the best for all organs (with respect to *DC* in Table 5), right lung and right kidney (with respect to mean-*HD* in Table 6) whereas Experiment 1 fares the best for the left lung, left kidney, and spleen, and Experiment 2 fares the best for the liver and thoraco-abdominal skin in Table 6. The competitive performance of Experiment 3 is in accordance to our intuition that if an image (say *A*) of a subject at a particular respiratory phase is included with it’s associated ground truth in the training of DL-D, then the delineation performance of DL-D for an image of the same subject at a different respiratory phase will be better compared to its performance when *A* is not used in training.

### 4.3. Exp. 4-6: Experiments which utilize (3D) images at multiple (>2) respiratory phases (EI, EE, and intermediate phases)

The problem of segmenting an object at any phase (Q) for a subject x given GT for one phase (P ≠ Q) for x, is also legitimate and practically very relevant. We conduct Exp. 4-6 where we assume that we have the ground truth for an organ of interest in an image of the test subject at a respiratory phase P which is different from the respiratory phase Q in which we are trying to delineate the organ.

Note that in the previous experiments, the bounding boxes for the delineation step were obtained from DL-R. However, in the incremental manner of predicting via DL-D, initially, the bounding boxes for delineation at a respiratory phase P1 closest to P comes from the enlarged tight fitting bounding box around the ground truth at P. The bounding boxes for the delineation at the next respiratory phase P2 closest to P1 comes from the enlarged tight fitting bounding boxes around the delineations at P1. This process continues recursively until we have the delineations for the organs in the image of the test subject at respiratory phase Q.

We present the performance of DL-D for the delineation of the seven organs in images of the 90 subjects at EE in an incremental manner (i.e., Exp. 4 where Q=EE and P=EI). Of these 90 subjects, 29 had one intermediate respiratory phase between EI and EE, 57 had 2 intermediate respiratory phases between EI and EE, and 4 had 3 intermediate respiratory phases between EI and EE. The number of intermediate respiratory phases is variable because of the different respiratory rates of the subjects [17] which affects the quality of the image which in turn determines how many respiratory phases within a respiratory cycle can be utilized to reliably reconstruct the image of the body region. The training (and validation) set consisted of images of the 89 (and 10) subjects at EI. The results in terms of average (and standard deviation) of Dice coefficients over all 90 test subjects for seven organs are shown in the second row (Experiment 4) of Table 7. The corresponding average and standard deviation of the mean-*HD* values are shown in Table 8.

**Table 7:** Performance (mean±SD: DC) of DL-D for delineation of left lung, right lung, left kidney, right kidney, liver, spleen, and thoraco-abdominal skin in an incremental manner in three different experiments. The highest Dice coefficient under a column has been highlighted.

|  |  |  |  |  |  |  |  |
| --- | --- | --- | --- | --- | --- | --- | --- |
| Experiment | Left lung | Right lung | Left kidney | Right kidney | Liver | Spleen | Thoraco-abdominal skin |
| Experiment 4 (90 EI to EE) | 0.937<br>±0.018 | **0.929**<br>**±0.040** | **0.820**<br>**±0.077** | 0.831<br>±0.086 | 0.898<br>±0.048 | 0.808<br>±0.073 | **0.949**<br>**±0.028** |
| Experiment 5 (90 EE to EI) | 0.922<br>$\pm 0.035$ | 0.923<br>$\pm 0.039$ | 0.814<br>$\pm 0.066$ | 0.830<br>$\pm 0.087$ | 0.897<br>$\pm 0.039$ | 0.811<br>$\pm 0.060$ | 0.936<br>$\pm 0.027$ |
| Experiment 6 (99 EI to EE) | **0.942**<br>**$\pm 0.026$** | 0.921<br>$\pm 0.052$ | 0.816<br>$\pm 0.070$ | **0.841**<br>**$\pm 0.097$** | **0.907**<br>**$\pm 0.020$** | **0.813**<br>**$\pm 0.071$** | 0.945<br>$\pm 0.026$ |

**Table 8:**
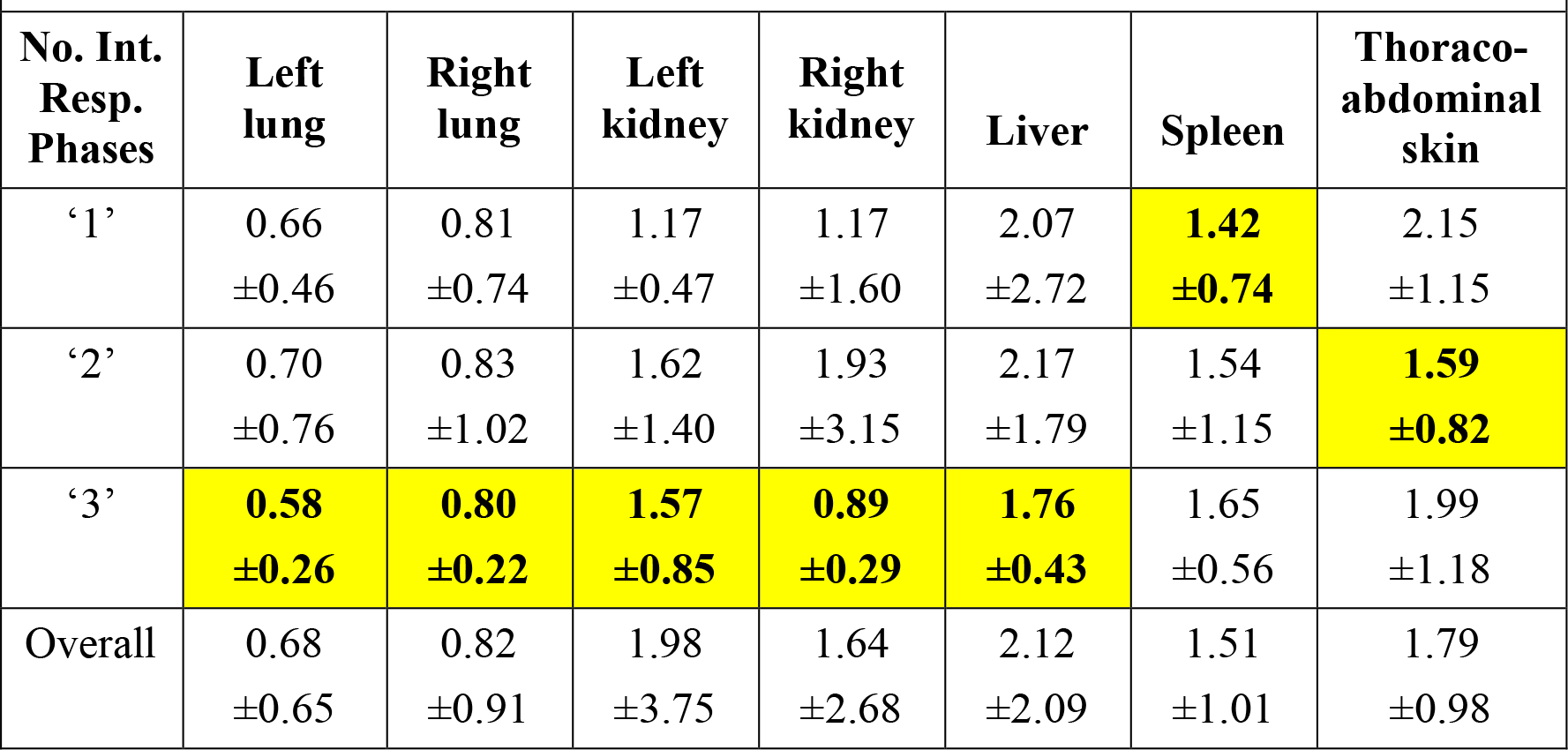
Performance of DL-D in terms of (mean±SD) mean-HD (mm) for Experiment 4. The 90 test images have been categorized in the first column based on the number of intermediate respiratory phases between EE and EI. For further details, please refer subsection 4.3.

In Experiment 5, we test the delineation of the seven organs on images of the 90 subjects at EI (Q=EI), using the ground truth of organs in the images of the aforesaid 90 subjects at EE (P=EE), in an incremental manner. The results are shown in (the third row as Experiment 5 of) Table 7 and Table 9.

**Table 9:** Performance of DL-D in terms of (mean±SD) mean-HD (mm) for Experiment 5. The 90 test images have been categorized in the first column based on the number of intermediate respiratory phases between EE and EI. For further details, please refer subsection 4.3.

| No. Int. Resp. Phases | Left lung | Right lung | Left kidney | Right kidney | Liver | Spleen | Thoraco-abdominal skin |
| --- | --- | --- | --- | --- | --- | --- | --- |
| ‘1’ | 0.51<br>$\pm 0.23$ | 0.76<br>$\pm 0.63$ | 1.48<br>$\pm 2.07$ | 1.23<br>$\pm 1.49$ | 2.13<br>$\pm 2.95$ | 1.80<br>$\pm 1.54$ | 2.83<br>$\pm 1.62$ |
| ‘2’ | 0.54<br>$\pm 0.23$ | 0.97<br>$\pm 1.43$ | 1.29<br>$\pm 0.83$ | 1.45<br>$\pm 1.68$ | 2.18<br>$\pm 1.96$ | **1.37**<br>**$\pm 0.63$** | 2.08<br>$\pm 1.08$ |
| ‘3’ | **0.39**<br>**$\pm 0.17$** | **0.55**<br>**$\pm 0.31$** | **0.89**<br>**$\pm 0.23$** | **0.81**<br>**$\pm 0.38$** | **1.50**<br>**$\pm 0.63$** | 1.40<br>$\pm 0.43$ | **1.49**<br>**$\pm 0.96$** |
| Overall | 0.52<br>$\pm 0.23$ | 1.12<br>$\pm 2.56$ | 1.33<br>$\pm 1.34$ | 1.57<br>$\pm 2.62$ | 2.13<br>$\pm 2.27$ | 1.51<br>$\pm 1.01$ | 2.29<br>$\pm 1.43$ |

In Experiment 6, we test the DL-D for the said seven organs on images of 99 subjects at EE (Q=EE) using the ground truth of the organs in the images of the aforesaid 99 subjects at EI (P=EI), in an incremental manner. Out of these 99 subjects, 9 did not have images at intermediate respiratory phases. Amongst the remaining 90 (=99-9) subjects, 28, 54, and 8 had images at 1, 2, and 3 intermediate respiratory phases, respectively. The results are shown in (the fourth row (Experiment 6) of) Table 7 and Table 10.

**Table 10:**
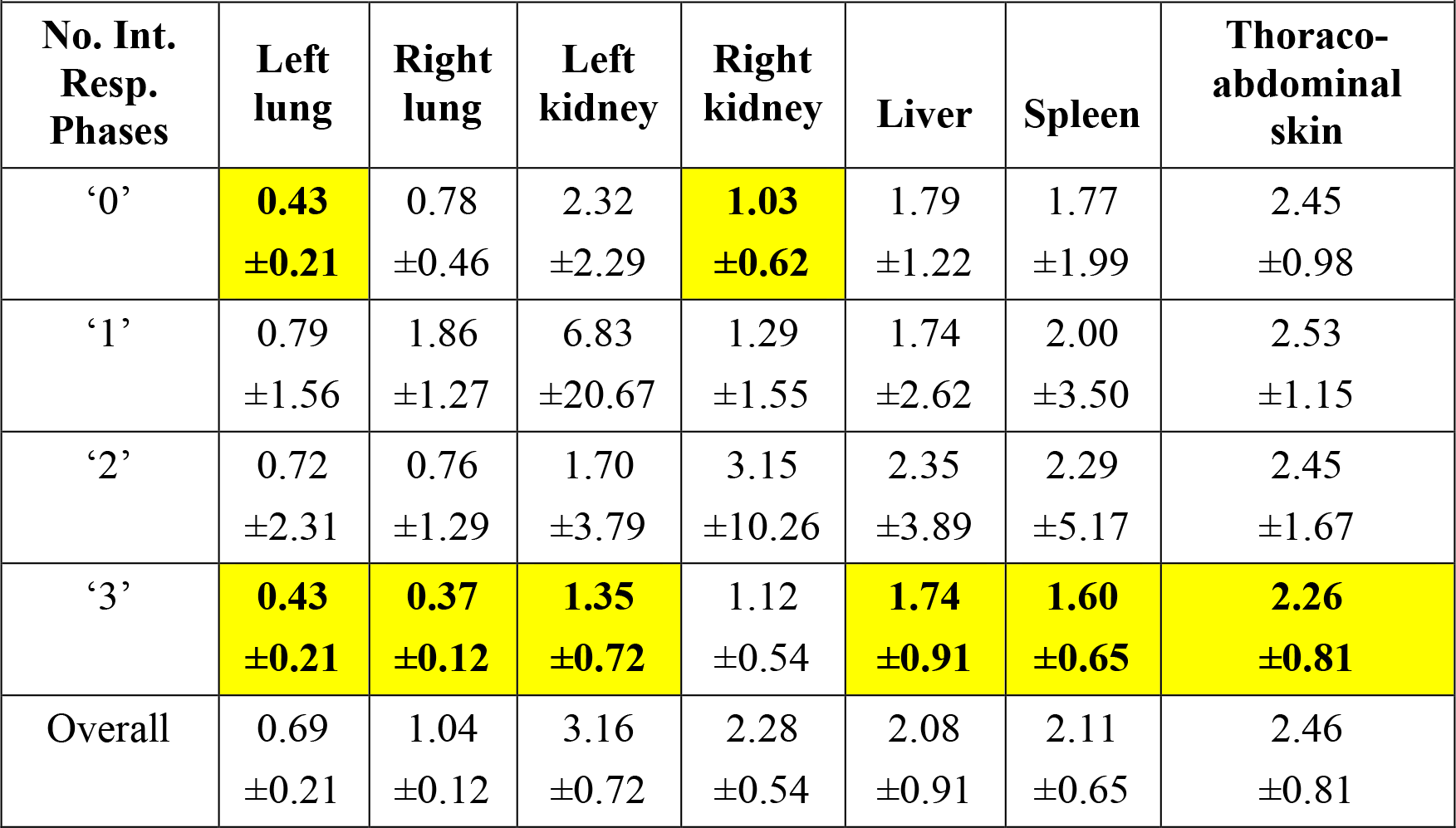
Performance of DL-D in terms of (mean±SD) mean-HD (mm) for Experiment 6. The 99 test images have been categorized in the first column based on the number of intermediate respiratory phases between EE and EI. The best performing case amongst ‘0’, ‘1’, ‘2’, and ‘3’ intermediate respiratory phases has been highlighted. For further details, please refer subsection 4.3.

Experiments 5 and 1 are similar in the sense that both try to delineate organs in images of 90 subjects at EI, and the same holds true for Experiments 4 and 2 at EE. The results of Experiment 5 in Table 7 are better by about 1% for the left kidney (0.74%), right kidney (0.24%), spleen (0.50%) and thoraco- abdominal skin (0.75%) compared to those of Experiment 1 in Table 5. Except for the right kidney, liver, and spleen, Experiment 4 yielded better results than Experiment 2 by about 1% to 3%. Experiments 6 and 3 are similar in the sense that both try to delineate organs in images of 99 subjects at EE. Experiment 6 yields inferior results by about 2% for the right lung, left kidney, and spleen compared to those of Experiment 3. For the remaining organs, Experiments 6 and 3 perform in a statistically similar fashion.

We notice that despite using additional information about the test image in Table 7, the results did not significantly improve compared to those in Table 5 with respect to *DC*. It seems that this is because the delineations at an intermediate respiratory phase are not sufficiently accurate to provide a correct bounding box for the delineation of the organs at the subsequent respiratory phase. We can expect this drawback to be minimal if the adjacent respiratory phases are closer to one another. For example, if we look at the results in Tables 8, 9, and 10 we notice that the mean-*HD* assumes the least value mostly at ‘3’ number of intermediate respiratory phases. This suggests that if we have images at a larger number of intermediate respiratory phases between EE and EI, then the incremental manner of testing DL-D would demonstrate better performance.

### 4.4 Exp. 7: Experiment on repeated scans of the same thoraco-abdominal region of the same subject

We reiterate that different dMRI scans even for the same body region and for the same subject need not have the same meaning of gray-level intensities for an organ [24]. Intensity standardization (IS) was developed to circumvent this problem [24]. In this experiment, we test whether our segmentation model can perform consistently in multiple dMRI scans of the same thoraco-abdominal region of a subject in repeated scans. We obtained 2 repeated dMRI scans of each of 5 subjects with a 5-minute gap between the two repeated scans and created auto-segmentations of both lungs for each subject. The mean and SD of DC values over the 5 scans are summarized in Table 11 as well as the *p* value comparing DCs between the two repeated scans.

**Table 11:** Performance of DL-D on repeated scans in Exp.7. For details refer to subsection 4.4.

|  |  |  |  |
| --- | --- | --- | --- |
| **Organ** | **1st scan** | **2nd scan** | **$p$-value** |
| Left lung | 0.948<br>$\pm 0.016$ | 0.900<br>$\pm 0.057$ | 0.078 |
| Right lung | 0.919<br>$\pm 0.064$ | 0.923<br>$\pm 0.034$ | 0.764 |

We observe that the performance of DL-D is similar in the two scans for the right lung (*p*=0.764 based on DC) and the left lung (*p*=0.078, α=0.05 based on DC). These results suggest that our segmentation set-up can perform consistently in multiple dMRI acquisitions of the same thoraco-abdominal region of a subject in most of the cases despite these acquisitions having different intensity meanings for an organ.

### 4.5 Exp. 8: Comparison with competing approaches

As mentioned previously, articles [15, 16] deal with the segmentation of the lungs only. In this experiment, we have therefore compared our work with these methods for the segmentation of just the two lungs.

We took the trained model of [15] as is for testing. As reported in [15], that model was trained using images of 36 subjects at EE. Of these 36 subjects, 29 belonged to the set of 99 subjects. In Table 12, we therefore show results of the method in [15] on images of the 70 (=99-29) subjects at EI and EE (Experiment 8). The corresponding average and standard deviation of mean-*HD* are shown in Table 13.

**Table 12:** Performance of a competing approach. [12] **for segmentation of left lung and right lung in terms of DC.**

|  |  |  |  |
| --- | --- | --- | --- |
| Category | Respiratory Phase | Left lung | Right lung |
| Experiment 8: 70 subjects | EI | 0.833<br>$\pm 0.046$ | 0.847<br>$\pm 0.040$ |
| | EE | 0.881<br>$\pm 0.098$ | 0.910<br>$\pm 0.051$ |

**Table 13:** Performance of a competing approach. [12] **for segmentation of left lung and right lung in terms of mean- HD.**

|  |  |  |  |
| --- | --- | --- | --- |
| Category | Respiratory Phase | Left lung | Right lung |
| Experiment 8: 70 subjects | EI | 2.62<br>$\pm 1.22$ | 2.94<br>$\pm 2.20$ |
| | EE | 2.23<br>$\pm 2.33$ | 2.21<br>$\pm 2.78$ |

Article [16] reports a Dice coefficient of 0.94-0.96 for the segmentation of the lungs in dMRI using atlas based registration methods. The dataset which they [16] have utilized is not the same as ours with regards to spatial resolution (2.81 X 2.81 X 4 *mm^3^*) and scanner and mode of acquisition (regulated breathing compared to our free-breathing acquisitions). The age of the patients have not been disclosed in [16].

## 5. Discussion and Conclusions

The results of Exp. 1, which are shown in the second row of Table 5, and the results of the last column of Table 2 are both obtained using images at EI in the training set as well as in the test set. The only difference between these two experiments is that the former (Experiment 1) utilized 189 subjects (∼100 images for training and validation) for evaluation while the latter utilized 95 subjects (∼80 images for training and validation) for evaluation. We notice that these two results are statistically similar. This observation suggests that at about 100 studies, the performance of DL-R and DL-D perhaps stabilize.

From the results in Table 5 and Table 12, we find that our proposed approach performs better by about 2% to 10% compared to [15]. As mentioned earlier, the approach of [15] is based on a 2D U-Net architecture whereas our architecture (ABCNet [21]) is based on an enhanced version of an encoder-decoder architecture. We think that judiciously designed sophisticated architecture like DL-R and DL-D modules can handle the challenges in segmentation from dMRI images better than networks such as in [15], since our set-up works better on unseen images.

The evaluations shown in this paper are based on a dMRI dataset from a single center. Gathering additional datasets from multiple clinical centers might help us to assess the robustness of the proposed approach. We believe that merging natural intelligence techniques with artificial intelligence techniques can have the potential to provide better segmentation performance. We will try to investigate the design of the hybrid intelligence framework for application to dMRI in patients with TIS as a future work.

For the sake of completeness, we have shown 6 slices for each organ where the ground truth for the organ is superimposed on the delineation by the proposed auto-segmentation set-up in Figure 4. In Figure 5, we have shown the 3D rendering of the prediction results of DL-D for the left lung, right lung, left kidney, right kidney, liver, and spleen for three subjects along with the 3D rendering of the corresponding ground truth of the said organs.

**Figure 4:**
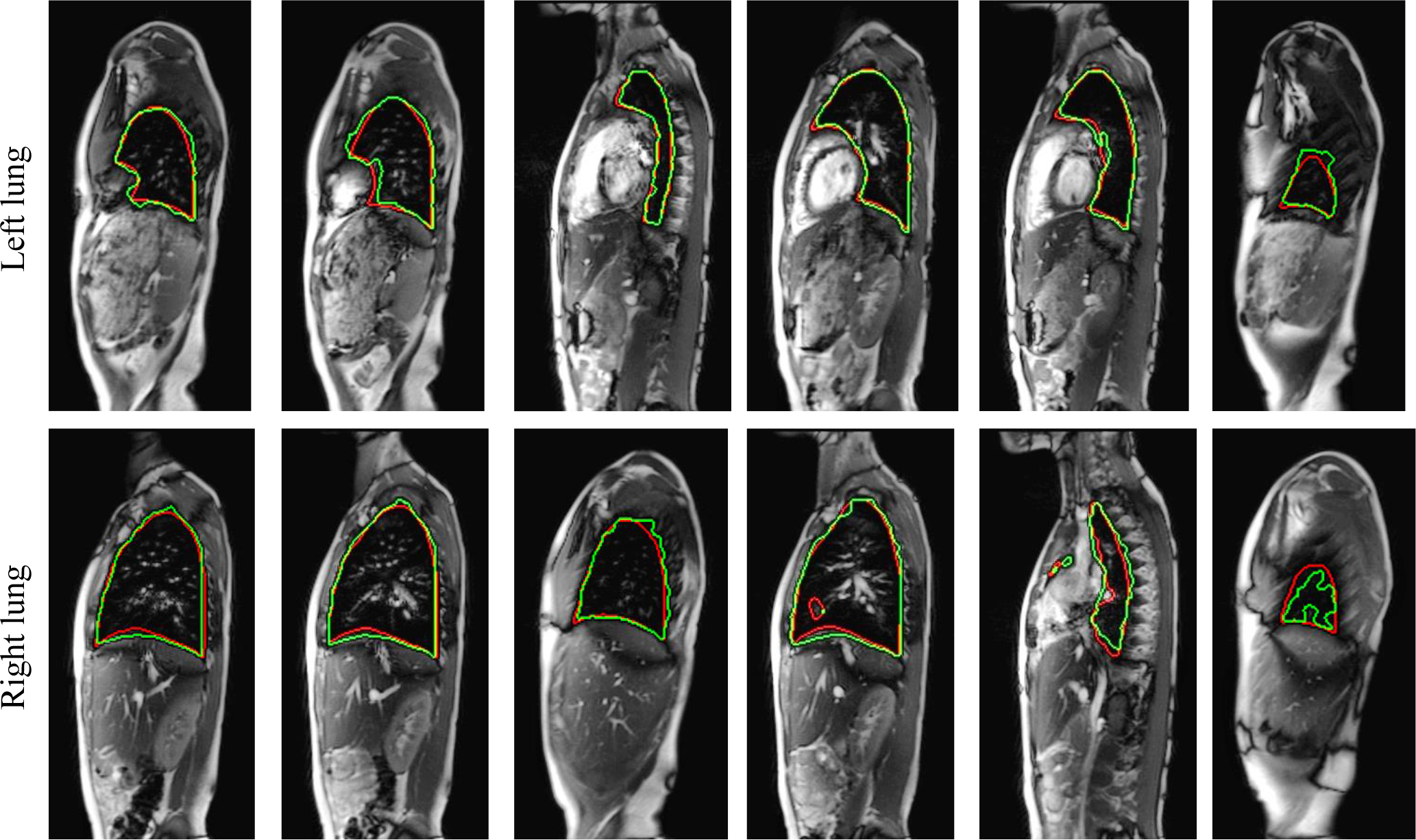
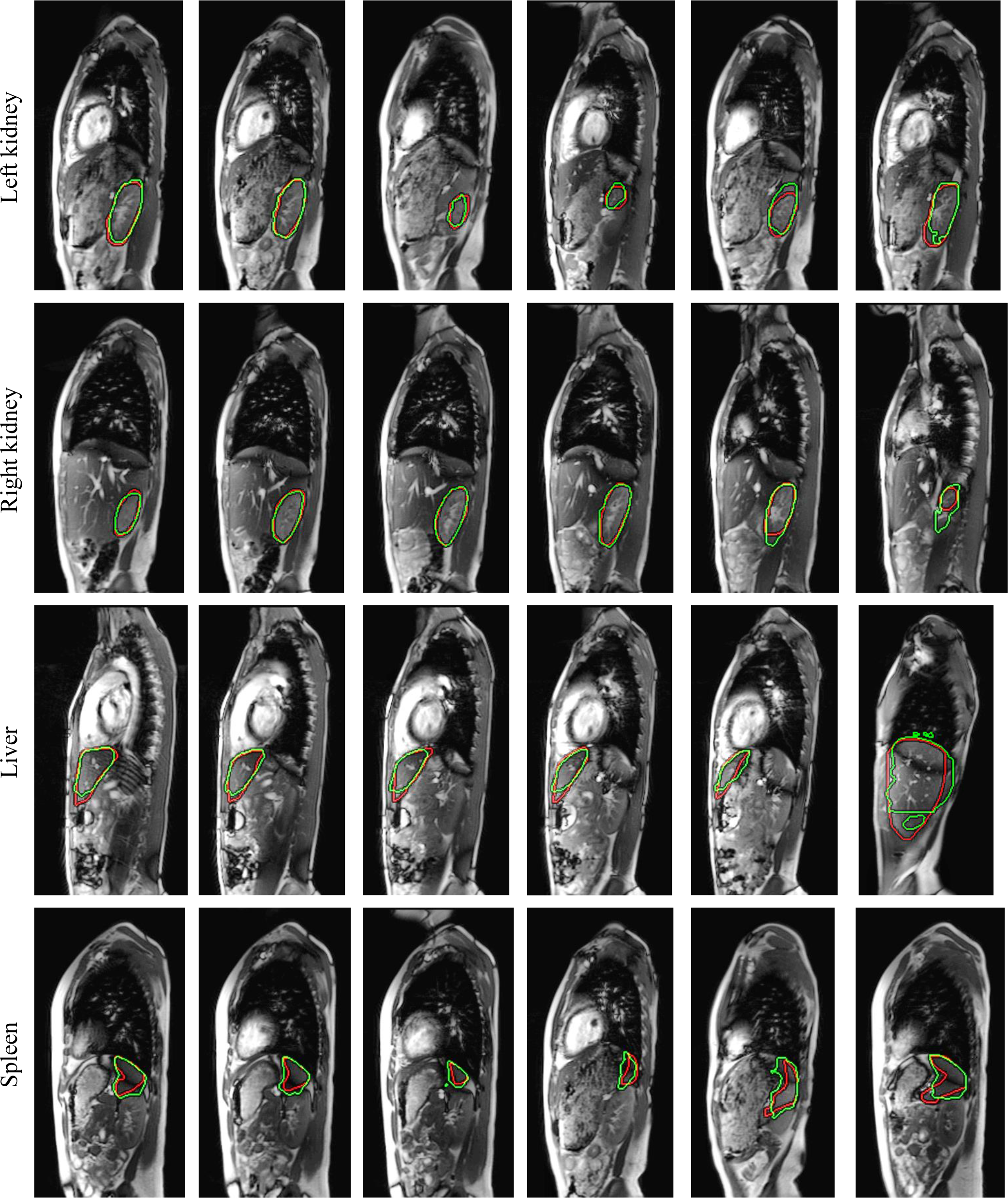
A row shows the delineations for an organ (top to bottom: left lung, right lung, left kidney, right kidney, liver, and spleen) by the proposed auto-segmentation algorithm (green) and by the expert human tracer (red) in six sagittal bright-blood dMRI slices. The last column shows those cases where the delineations by the proposal are relatively more off compared to the ground truth.

**Figure 5:**
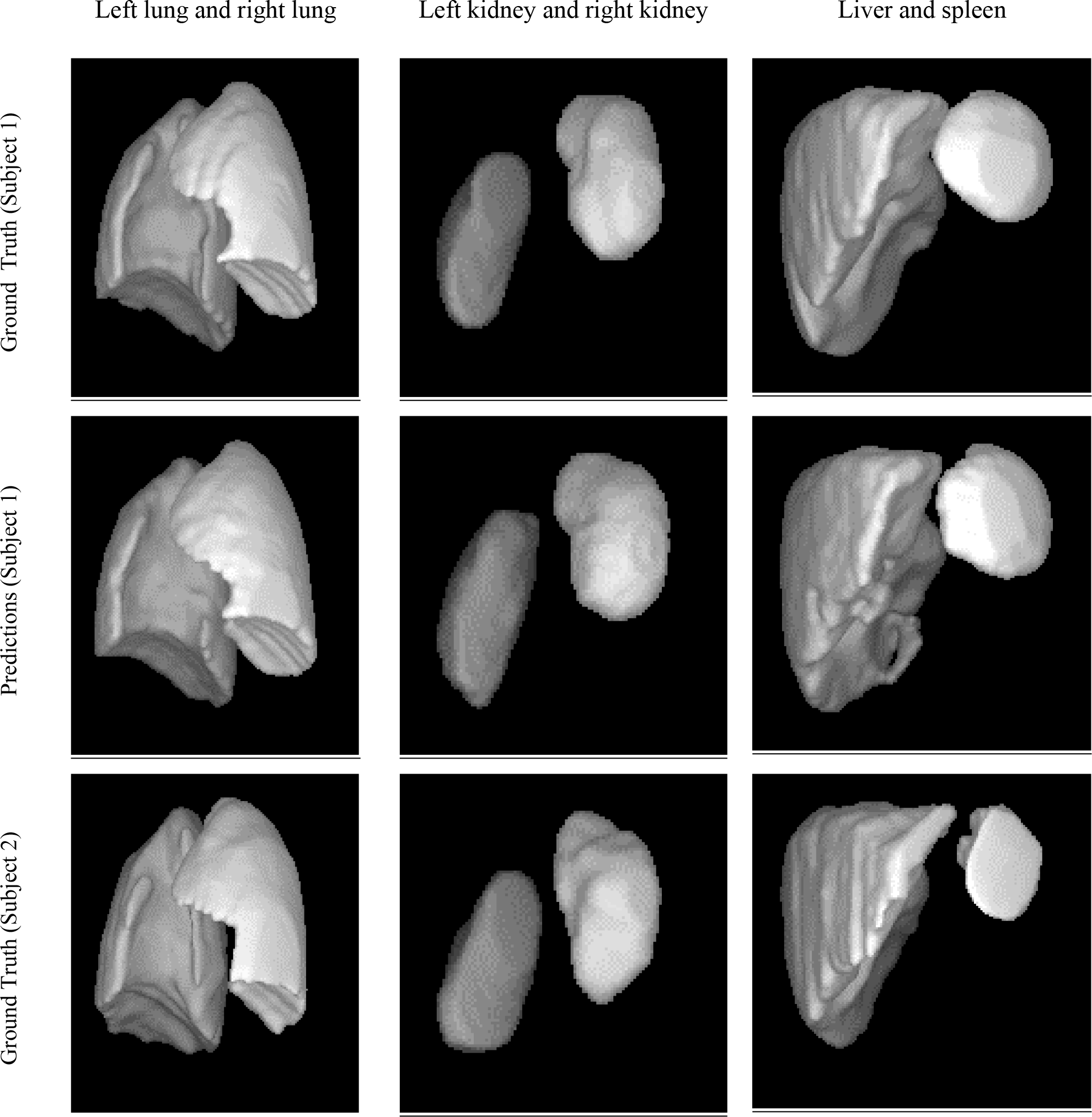
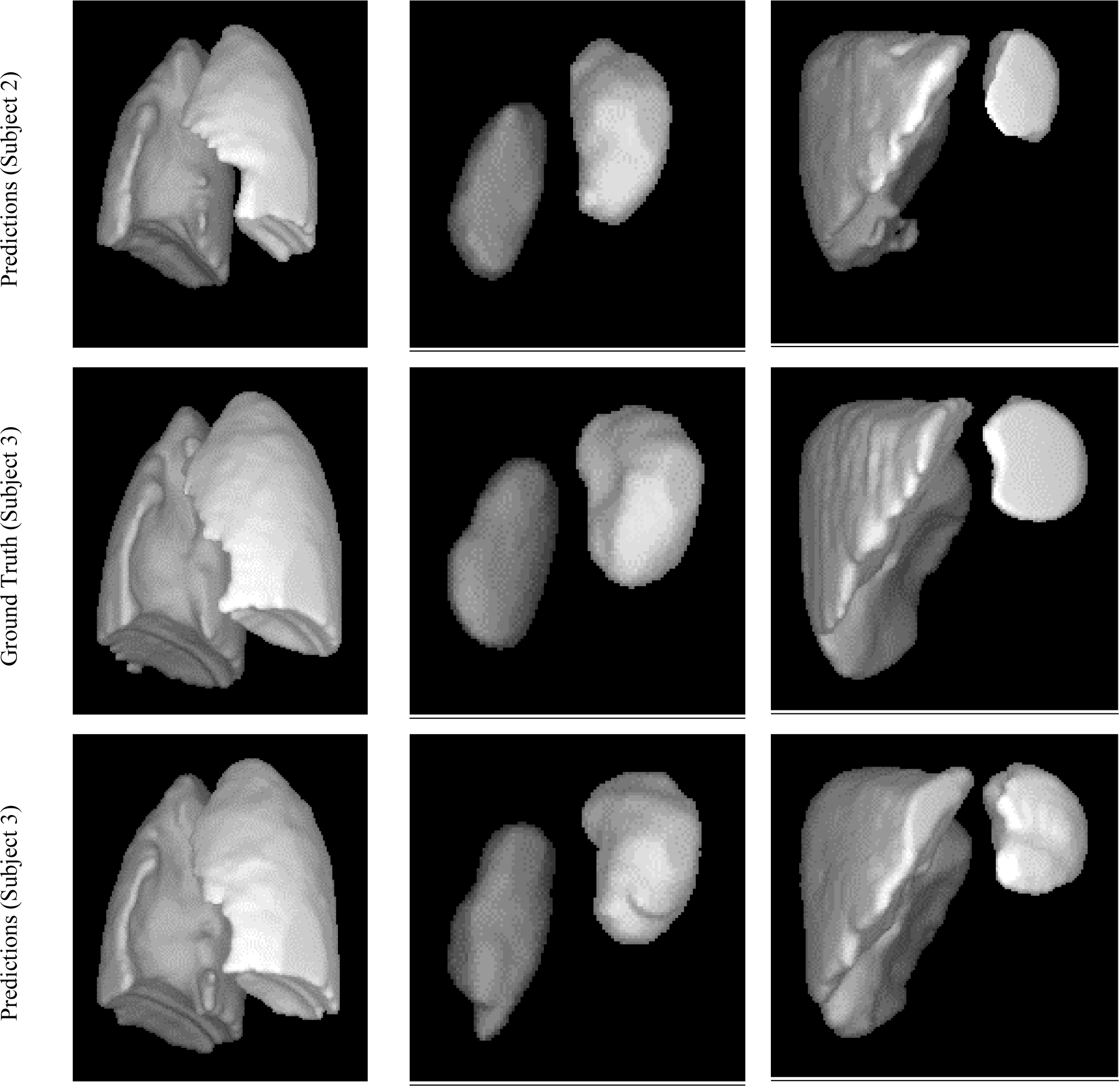
3D rendering of ground truth (1^st^ row) and corresponding predictions (2^nd^ row) of a subject for left lung and right lung (1^st^ column), liver and spleen (2^nd^ column), and left kidney and right kidney (3^rd^ column). The illustration has been repeated for a second subject in the 3^rd^ row (ground truth) and 4^th^ row (predictions) and for a third subject in the 5^th^ row (ground truth) and 6^th^ row (predictions).

In this paper, we have developed an auto-segmentation setup for delineation of the thoraco-abdominal organs in dynamic magnetic resonance imaging (dMRI) images of pediatric subjects. We have implemented the segmentation in two steps: a recognition step and a delineation step. The DL-R is reasonably able to localize the organs given that the dMRI images are challenging to handle compared to other imaging modalities. For the delineation of the organs, we compared two AI approaches: ABCNet [21] and U-Net [15]. The delineation results for the lungs, kidneys, liver, and spleen by ABCNet from dMRI sagittal image acquisitions of the thoraco-abdominal region of (near-)normal healthy subjects are excellent, in light of the extreme difficulty of segmentation of these objects in dMRI images. We are further investigating this system for segmentation of the thoraco-abdominal organs in dMRI images of patients with thoracic insufficiency syndrome.

## Data Availability

All data produced in the present work are contained in the manuscript.
All data produced in the present study are available upon reasonable request to the authors

## Acknowledgements

This research was supported by a grant from the National Institutes of Health R01HL150147.

